# Depression at the intersection of race/ethnicity, sex/gender, and sexual orientation in a nationally representative sample of US adults: A design-weighted MAIHDA

**DOI:** 10.1101/2023.04.13.23288529

**Authors:** F. Hunter McGuire, Ariel L. Beccia, JaNiene Peoples, Matthew R. Williams, Megan S. Schuler, Alexis E. Duncan

## Abstract

This study examined how race/ethnicity, sex/gender, and sexual orientation intersect to socially pattern depression among US adults. We used repeated, cross-sectional data from the 2015-2020 National Survey on Drug Use and Health (NSDUH; n=234,772) to conduct design-weighted multilevel analysis of individual heterogeneity and discriminatory accuracy (MAIHDA) for two outcomes: past-year and lifetime major depressive episode (MDE). With 42 intersectional groups constructed from seven race/ethnicity, two sex/gender, and three sexual orientation categories, we estimated group-specific prevalence and excess/reduced prevalence attributable to intersectional effects (i.e., two-way or higher interactions between identity variables). Models revealed heterogeneity between intersectional groups, with prevalence estimates ranging from 3.4–31.4% (past-year) and 6.7–47.4% (lifetime). Model main effects indicated that people who were Multiracial, White, women, gay/lesbian, or bisexual had greater odds of MDE. Additive effects of race/ethnicity, sex/gender, and sexual orientation explained most between-group variance; however, approximately 3% (past-year) and 12% (lifetime) were attributable to intersectional effects, with some groups experiencing excess/reduced prevalence. For both outcomes, sexual orientation main effects (42.9–54.0%) explained a greater proportion of between-group variance relative to race/ethnicity (10.0–17.1%) and sex/gender (7.5–7.9%). Notably, we extend MAIHDA to calculate nationally representative estimates to open future opportunities to quantify intersectionality with complex sample survey data.

## INTRODUCTION

Depression is a serious psychiatric disorder associated with adverse health outcomes and comorbidities, including cognitive impairments,^1^ substance use disorders,^2^ excess mortality,^3^ and suicidality.^4^ In recent years, the prevalence of depression has increased considerably among US adults.^5^ The economic burden of depression has concurrently increased from an inflation-adjusted $227 billion in 2010 to $326 billion in 2018,^5^ and few US adults with depression report utilizing mental health services.^6,7^ Therefore, identifying subpopulations experiencing a disproportionate burden of depression is critical for surveillance guiding prevention and intervention efforts to counteract depression-related costs, morbidity, and mortality.

### Social epidemiology of depression by race/ethnicity, sex/gender, and sexual orientation

Prior research has documented depression prevalence differences comparing US adult subgroups defined by race/ethnicity,^7–14^ sex/gender,^8,15–18^ and sexual orientation.^19–22^ We use the term “sex/gender” to acknowledge the conflation of sex assigned at birth with gender identity arising from measurement decisions affecting the current study’s data and, where relevant, data from prior studies.^23^ Building upon community knowledge and activism, a growing evidence base is revealing how these differences are inequities attributable to systems of oppression (e.g., structural sexism, racism, and heterosexism) structuring power relations in US society and how these power relations are incorporated into and reproduced by social, familial, economic, and political systems.^24–26^ Overall, this structure maintains societal conditions where White people (vs. people of color), boys/men (vs. girls/women and other marginalized sex/gender groups), and heterosexual people (vs. sexual minority people) have greater accumulations of and access to power.

Overall, findings for racial/ethnic inequities in depression among US adults are mixed concerning strength and direction. Studies have generally found that non-Hispanic White adults have increased odds of depression compared to adults who are non-Hispanic Black,^7–9,12–14^ non-Hispanic Asian or Pacific Islander,^8^ and Hispanic.^8,9^ However, other data suggest the prevalence of past-year major depressive episode (MDE) is highest among Puerto Ricans and lowest among Asian subgroups (i.e., Chinese, Filipino, and Vietnamese Americans), while other Hispanic subgroups (i.e., Mexican and Cuban Americans) had similar prevalence of past-year MDE compared to non-Hispanic White adults.^11^ Another study found that, among US older adults, those who identified as Hispanic, Black, Asian Indian, Filipino, Native Hawaiian or Pacific Islander, or Multiracial had elevated odds of screening positive for depression relative to non-Hispanic White people.^10^ For studies investigating differences across binary categories of sex/gender (i.e., men vs. women), findings consistently estimate greater prevalence of depression among women.^8,15–18^ Prior research also indicates that sexual minority people (e.g., those who identify as gay, lesbian, or bisexual and/or who have same or multiple gender partners or attractions) generally have an elevated prevalence of depression relative to heterosexual people.^19–22^

### Intersectionality as a conceptual and analytical tool

Accumulating evidence suggests that privilege and/or marginalization across systems of oppression may combine in complex ways to produce population-level health inequities.^24–26^ Originating in Black feminist scholarship,^27,28^ intersectionality is a theoretical orientation that emphasizes the interwoven ways in which identities at the individual level (e.g., race/ethnicity, sex/gender, sexual orientation) interact with interlocking systems of oppression at the macro-level (e.g., racism, sexism, heterosexism) to produce a complex social patterning of health and wellbeing. Intersectionality stresses that identities are mutually constitutive and interactions between them are non-additive.^29,30^ As such, systems of oppression are interwoven in a way that, for example, the experience of a Black woman’s oppression is not simply the experience of racial oppression paired with the experience of sex/gender-based oppression – it can take on a form that is unique to the intersectional position.

Intersectionality has in recent years been applied in the analysis and interpretation of epidemiological research.^31^ Despite prior evidence highlighting inequities across unidimensional identity measures, the extent that depression among US adults is patterned at the intersection of race/ethnicity, sex/gender, and sexual orientation remains poorly understood, although some prior studies using US nationally representative data have examined the intersection of two of these identities. For example, a study considering the intersection of sex/gender and sexual orientation found that bisexual women had the highest prevalence of past-year and lifetime MDE,^20^ while another study investigating intersectionality of race/ethnicity and sexual orientation found that sexual minority groups across race/ethnicity (i.e., White, Black, Hispanic) had similarly high prevalence of past-year MDE.^32^

A recently developed analytic approach for intersectional analyses is multilevel analysis of individual heterogeneity and discriminatory accuracy (MAIHDA),^33^ which uses multilevel modeling to estimate health outcome prevalence and intersectional effects across large numbers of intersectional groups. Intersectional effects describe how group-specific prevalence differs from expectations based on independent, additive contributions of each identity variable in the absence of interactions. A prior MAIDHA study investigated depression symptom trajectories among US adolescents and young adults and documented heterogeneity at the intersection of race/ethnicity, sex/gender, immigration status, and income.^34^ However, this analysis did not include sexual orientation and was limited to younger age groups, so how sexual orientation, in combination with race/ethnicity and sex/gender, may structure the social patterning of depression across a wide age range of US adults remains unknown. This is particularly important given prior evidence documenting large sexual orientation-based inequities in depression.^19–22^

### Current study

Using combined data from the National Survey on Drug Use and Health (NSDUH) 2015-2020, the current study investigates how race/ethnicity, sex/gender, and sexual orientation (as imperfect proxies for exposure to structural racism, sexism, and heterosexism) intersect to structure the patterning of past-year and lifetime MDE among US adults. Under an intersectional framework^27,28^ and building on the formative work of Evans and colleagues^33^ and Merlo,^35^ we implement a design-weighted MAIHDA to estimate the prevalence of past-year and lifetime MDE. Our approach extends prior MAIHDA applications by generating nationally representative estimates through the incorporation of complex sample survey weights. We additionally estimate intersectional group-specific excess/reduced prevalence due to interaction, defined here as the extent to which the prevalence of MDE for a given intersectional group is greater (or lesser) than what would be expected based on the additive effects (i.e., addition of main effects) of its constituent race/ethnicity, sex/gender, and sexual orientation categories.

## METHODS

### Data source and analytic sample

Data came from six repeated, cross-sectional waves of NSDUH collected from 2015 to 2020. NSDUH uses a multi-stage probability sampling design to collect data representative of non-institutionalized US residents aged 12 years and older. Data from 2015 to March 2020 were collected via computer-assisted in-person interviewing. Due to the COVID-19 pandemic, data collection was paused from mid-March to September 2020 with web-based data collection implemented from October to December 2020. Additional information on sampling and data collection can be found elsewhere.^36^ Data were de-identified and publicly available through the Substance Abuse and Mental Health Services Administration (SAMHSA).^37^ No additional ethics review was required.

From the overall dataset (n=315,661), we identified an analytic sample for each outcome. All participants had complete data for race/ethnicity and sex/gender, but we excluded those younger than 18 (n=73,986) as they were not asked about sexual orientation. After excluding those with missing data for lifetime MDE or sexual orientation (n=6,953), the total sample size was 234,772 for lifetime MDE models. Since additional participants (n=348) were missing on past-year MDE, the sample size for past-year MDE models was 234,374.

### Measures

#### Outcome measures: Lifetime and past-year major depressive episode (MDE)

Participants aged 18 or older completed a depression module using major depressive disorder criteria from the Diagnostic and Statistical Manual of Mental Disorders 5th Edition (DSM-5).^38^ Lifetime MDE was defined as reporting depressed mood and/or anhedonia and at least four or more of the following symptoms nearly every day in at least one 2-week period: (1) changes in appetite or weight, (2) sleep problems, (3) restlessness, (4) fatigue, (5) trouble concentrating, (6) feeling worthless, and (7) suicidal ideation. To measure past-year MDE, participants with a lifetime MDE were asked if these symptoms occurred during at least one 2-week period in the past 12-months.

### Race/ethnicity

Participants reported their race by choosing one or more options from: “White,” “Black or African American,” “American Indian or Alaska Native,” “Native Hawaiian,” “Guamanian or Chamorro,” “Samoan,” “Other Pacific Islander,” “Asian (including: Asian Indian, Chinese, Filipino, Japanese, Korean, And Vietnamese),” and “Other (Specify).” For ethnicity, participants were asked “Are you of Hispanic, Latino, or Spanish origin or descent?” with options “Yes” or “No,” and in this paper we refer to those who responded “Yes” with the gender-neutral term “Hispanic/Latine.” Race/ethnicity was categorized into seven groups: Hispanic/Latine, Non-Hispanic/Latine (NHL) Asian, NHL Black or African American, NHL Native American or Alaska Native, NHL Native Hawaiian or Pacific Islander, NHL Multiracial, or NHL White. Participants were coded as NHL Multiracial if they reported an NHL ethnicity and selected more than one race. Participants were coded as NHL Native Hawaiian or Pacific Islander if they had an NHL ethnicity and selected a race of Native Hawaiian, Guamanian or Chamorro, Samoan, or Other Pacific Islander. For brevity, we refer to racial groups without the “NHL” qualifier in this paper.

#### Sex/gender

Participants were not asked to self-report their sex assigned at birth or gender identity. NSDUH interviewers were instructed to “Record respondent’s gender” as either “Male” or “Female” and to ask participants to self-report “only if not obvious,” and the variable name (“IRSEX”) and label (“Gender”) in the codebook conflates sex with gender. We refer to this measure as “sex/gender” and use the terms “man” and “woman.”

#### Sexual orientation

Participants were asked to self-report their sexual orientation with “Which one of the following do you consider yourself to be?” with options of “Heterosexual, that is, straight,” “Gay”, or “Bisexual.” Participants who were interviewer-assigned as “Female” were given the option of “Lesbian or Gay” instead of “Gay.”

#### Intersectional groups

We sorted participants into intersectional groups using 42 mutually exclusive combinations of the seven race/ethnicity, two sex/gender, and three sexual orientation categories. Sample sizes for intersectional groups ranged from 16 to 69,450 (Table S1 in Supplementary Materials), and all groups were retained for analysis and interpretation.

#### Covariate

All models were adjusted for age category (18-25 [reference]), 26-34, 35-49, 50+) given prior evidence indicating that younger, relative to older, people have both a greater likelihood of experiencing depression^5^ and identifying as gay, lesbian, or bisexual^39^ and a lower likelihood of identifying as White.^40^

### Data analysis

#### Multilevel analysis of individual heterogeneity and discriminatory accuracy (MAIHDA)

The current study used Bayesian statistical methods to estimate the intersectional group-specific prevalence of each outcome using two-level logistic models. Individuals (level 1) were nested within intersectional groups (level 2). Model equations are available in Supplementary Materials. For each outcome, Model 1 was an age-adjusted intersectional model (i.e., age category as the only level 1 covariate) with random intercepts for intersectional groups. A series of intermediary models separately adjusted for race/ethnicity (Model 2a), sex/gender (Model 2b), and sexual orientation (Model 2c) as level 1 covariates to assess the relative importance of each identity variable in explaining group-level outcome variance. Finally, we estimated an age-adjusted intersectional interaction model (Model 3) with age category and all three identity variables as level 1 covariates.

We calculated measures of sample-level and group-specific intersectionality effects. Sample-level measures included the variance partition coefficient (VPC) and the proportional change in variance (PCV). As a measure of discriminatory accuracy, the VPC represents the proportion of model variance attributable to differences between intersectional groups (i.e., the ability of intersectional groups to distinguish cases from non-cases of a given outcome). Relative to Model 1, the PCV measures the proportion of group-level outcome variance attributable to the additive effects (i.e., addition of main effects) of identity variables. Higher PCV values in Models 2a-c indicates higher incremental discriminatory accuracy (i.e., greater relevance of the identity variable in explaining group-level outcome variance). The Model 3 PCV shows the proportion of group-level variance from Model 1 attributable to additive effects of all identity variables considered simultaneously.

Group-specific intersectionality effects were estimated with (1) model-predicted age-adjusted prevalence estimates (calculated from Model 1 by combining estimates of each group-specific intercept with the overall intercept and converting from log-odds to probability scale) and (2) excess/reduced prevalence due to intersectional effects (calculated by isolating group-specific intercept estimates from Model 3 which represent residual variance across intersectional groups unexplained by level 1 covariates). Finally, we obtained odds ratios (ORs) with 95% credible intervals (CIs) in Models 2a-c and Model 3 to characterize the posterior distribution of each estimate. Contrary to frequentist confidence intervals, Bayesian 95% credible intervals are interpreted as a 95% probability that the interval contains the population parameter value given the data, model, and priors.^41^

#### Design-weighted estimation procedures

Models were fit with Markov chain Monte Carlo (MCMC) with a Bernoulli outcome distribution and logit link. Non-informative priors for all parameters and four Markov chains were specified. Chains in Models 1 and 2a-c were run for 5,000 iterations including 1,000 warmup iterations (total post-warmup iterations = 16,000). Due to increased model complexity, chains in Model 3 were run for 10,000 iterations including 2,500 warmup iterations (total post-warmup iterations = 30,000). Model convergence was evaluated with Gelman-Rubin r-hat diagnostics and visual inspection of trace and autocorrelation plots.^42^

Our approach incorporated NSDUH design weights to generate US nationally representative estimates and account for the complex sample survey design.^43^ First, we divided the person-level weights by six to adjust for the six years of repeated cross-sectional data in the current study. We then scaled the person-level weights to ensure compatibility with MCMC model weights. Using the “survey” package,^44^ we created a survey design object with the weights and overall dataset as inputs and subset this object to identify the analytic subpopulation for each outcome variable. Finally, we used the “csSampling”^45^ package and author-defined functions to generate design-weighted pseudo-posterior distributions for each parameter. All analyses were conducted in R 4.2.2.

## RESULTS

### Sample characteristics

Frequencies and design-weighted percentages for model variables are presented in Table 1. Most participants were White (64.4%), women (51.5%), and heterosexual (94.7%). A plurality of participants were age 50 or older (45.6%). For outcome variables, 7.4% had a past-year MDE while 14.2% had a lifetime MDE.

**Table 1.** Demographic characteristics, National Survey on Drug Use and Health (NSDUH) 2015-2020 (n=234,772)

| Variables | n (weighted %) |
| --- | --- |
| **Dimensions of social identity/position** |  |
| Race/ethnicity |  |
| Asian | 11,014 (5.4%) |
| Black or African-American | 28,500 (11.8%) |
| Hispanic or Latine | 38,933 (15.8%) |
| Native American or Alaska Native | 3,162 (0.5%) |
| Native Hawaiian or Pacific Islander | 1,119 (0.4%) |
| Multiracial | 7,774 (1.7%) |
| White | 144,220 (64.4%) |
| Sex/gender |  |
| Woman | 125,591 (51.5%) |
| Man | 109,131 (48.5%) |
| Sexual orientation |  |
| Heterosexual | 217,207 (94.7%) |
| Gay or Lesbian | 5,314 (2.0%) |
| Bisexual | 12,201 (3.3%) |
| **Covariate** |  |
| Age category |  |
| 18-25 | 75,591 (13.8%) |
| 26-34 | 48,242 (15.9%) |
| 35-49 | 62,110 (24.6%) |
| 50+ | 48,779 (45.6%) |
| **Outcomes** |  |
| Past-year major depressive episode<sup>1</sup> | 22,224 (7.4%) |
| Lifetime major depressive episode | 39,446 (14.2%) |
| **NSDUH data collection year** |  |
| 2015 | 42,547 (16.5%) |
| 2016 | 41,626 (16.6%) |
| 2017 | 41,534 (16.8%) |
| 2018 | 41,837 (16.8%) |
| 2019 | 41,547 (16.9%) |
| 2020 | 25,631 (16.4%) |

<sup>1</sup> Missing data for 393 participants.
*Note:* Percentages are design-weighted to account for NSDUH complex sample survey weights.

### Design-weighted MAIHDA

Results from design-weighted MAIDHA of past-year MDE and lifetime MDE are shown in Tables 2 and 3, respectively. Prevalence estimates with 95% CIs are displayed in Figures 1 and 2. We found evidence of substantial heterogeneity in MDE prevalence across intersectional groups. Groups with greatest prevalence of MDE included Multiracial bisexual women (past-year: 31.4%; lifetime: 45.8%), White bisexual women (past-year: 30.7%; lifetime: 47.4%), and Multiracial bisexual men (past-year: 22.6%; lifetime: 30.8%), while groups with lowest prevalence of MDE were Asian heterosexual men (past-year: 3.4%; lifetime: 7.6%), Black heterosexual men (past-year: 4.0%; lifetime: 6.7%), and Hispanic/Latine heterosexual men (past-year: 4.1%; lifetime: 7.9%).

**Table 2.**
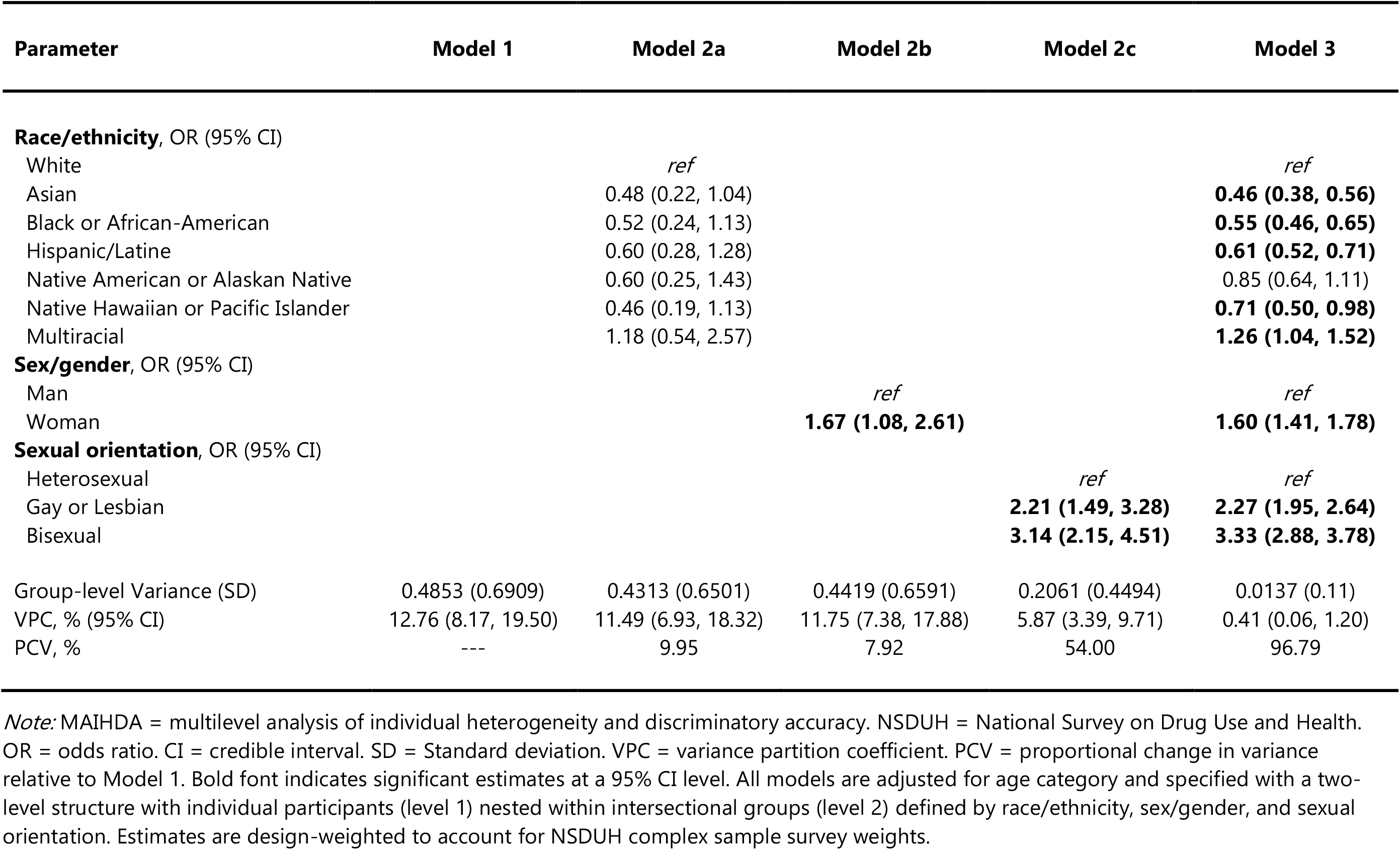
Design-weighted MAIHDA of past-year major depressive episode among US adults, NSDUH 2015-2020 (n=234,374)

| Parameter | Model 1 | Model 2a | Model 2b | Model 2c | Model 3 |
| --- | --- | --- | --- | --- | --- |
| **Race/ethnicity, OR (95% CI)** |  |  |  |  |  |
| White |  | *ref* |  |  | *ref* |
| Asian |  | 0.48 (0.22, 1.04) |  |  | **0.46 (0.38, 0.56)** |
| Black or African-American |  | 0.52 (0.24, 1.13) |  |  | **0.55 (0.46, 0.65)** |
| Hispanic/Latine |  | 0.60 (0.28, 1.28) |  |  | **0.61 (0.52, 0.71)** |
| Native American or Alaskan Native |  | 0.60 (0.25, 1.43) |  |  | 0.85 (0.64, 1.11) |
| Native Hawaiian or Pacific Islander |  | 0.46 (0.19, 1.13) |  |  | **0.71 (0.50, 0.98)** |
| Multiracial |  | 1.18 (0.54, 2.57) |  |  | **1.26 (1.04, 1.52)** |
| **Sex/gender, OR (95% CI)** |  |  |  |  |  |
| Man |  |  | *ref* |  | *ref* |
| Woman |  |  | **1.67 (1.08, 2.61)** |  | **1.60 (1.41, 1.78)** |
| **Sexual orientation, OR (95% CI)** |  |  |  |  |  |
| Heterosexual |  |  |  | *ref* | *ref* |
| Gay or Lesbian |  |  |  | **2.21 (1.49, 3.28)** | **2.27 (1.95, 2.64)** |
| Bisexual |  |  |  | **3.14 (2.15, 4.51)** | **3.33 (2.88, 3.78)** |
| Group-level Variance (SD) | 0.4853 (0.6909) | 0.4313 (0.6501) | 0.4419 (0.6591) | 0.2061 (0.4494) | 0.0137 (0.11) |
| VPC, % (95% CI) | 12.76 (8.17, 19.50) | 11.49 (6.93, 18.32) | 11.75 (7.38, 17.88) | 5.87 (3.39, 9.71) | 0.41 (0.06, 1.20) |
| PCV, % | --- | 9.95 | 7.92 | 54.00 | 96.79 |

*Note:* MAIHDA = multilevel analysis of individual heterogeneity and discriminatory accuracy. NSDUH = National Survey on Drug Use and Health. OR = odds ratio. CI = credible interval. SD = Standard deviation. VPC = variance partition coefficient. PCV = proportional change in variance relative to Model 1. Bold font indicates significant estimates at a 95% CI level. All models are adjusted for age category and specified with a two-level structure with individual participants (level 1) nested within intersectional groups (level 2) defined by race/ethnicity, sex/gender, and sexual orientation. Estimates are design-weighted to account for NSDUH complex sample survey weights.

**Table 3.** Design-weighted MAIHDA of lifetime major depressive episode among US adults, NSDUH 2015-2020 (n=234,722)

| Parameter | Model 1 | Model 2a | Model 2b | Model 2c | Model 3 |
| --- | --- | --- | --- | --- | --- |
| **Race/ethnicity, OR (95% CI)** |  |  |  |  |  |
| White |  | *ref* |  |  | *ref* |
| Asian |  | **0.41 (0.20, 0.85)** |  |  | **0.42 (0.31, 0.56)** |
| Black or African-American |  | **0.43 (0.21, 0.90)** |  |  | **0.44 (0.34, 0.59)** |
| Hispanic/Latine |  | 0.55 (0.27, 1.12) |  |  | **0.55 (0.42, 0.73)** |
| Native American or Alaskan Native |  | 0.50 (0.23, 1.10) |  |  | **0.63 (0.43, 0.90)** |
| Native Hawaiian or Pacific Islander |  | **0.39 (0.17, 0.91)** |  |  | **0.58 (0.38, 0.86)** |
| Multiracial |  | 1.01 (0.49, 2.08) |  |  | 1.08 (0.79, 1.45) |
| **Sex/gender, OR (95% CI)** |  |  |  |  |  |
| Man |  |  | *ref* |  | *ref* |
| Woman |  |  | **1.65 (1.07, 2.55)** |  | **1.51 (1.27, 1.81)** |
| **Sexual orientation, OR (95% CI)** |  |  |  |  |  |
| Heterosexual |  |  |  | *ref* | *ref* |
| Gay or Lesbian |  |  |  | **2.14 (1.40, 3.23)** | **2.19 (1.75, 2.70)** |
| Bisexual |  |  |  | **2.72 (1.79, 4.06)** | **2.78 (2.24, 3.41)** |
| Group-level Variance (SD) | 0.4781 (0.686) | 0.3866 (0.6158) | 0.4377 (0.6563) | 0.2569 (0.5018) | 0.0514 (0.2209) |
| VPC, % (95% CI) | 12.60 (8.08, 18.93) | 10.44 (6.31, 16.62) | 11.66 (7.39, 17.71) | 7.20 (4.24, 11.81) | 1.53 (0.56, 3.34) |
| PCV, % | --- | 17.14 | 7.46 | 42.86 | 87.86 |

*Note:* MAIHDA = multilevel analysis of individual heterogeneity and discriminatory accuracy. NSDUH = National Survey on Drug Use and Health. OR = odds ratio. CI = credible interval. SD = Standard deviation. VPC = variance partition coefficient. PCV = proportional change in variance relative to Model 1. Bold font indicates significant estimates at a 95% CI level. All models are adjusted for age category and specified with a two-level structure with individual participants (level 1) nested within intersectional groups (level 2) defined by race/ethnicity, sex/gender, and sexual orientation. Estimates are design-weighted to account for NSDUH complex sample survey weights.

**Figure 1.**
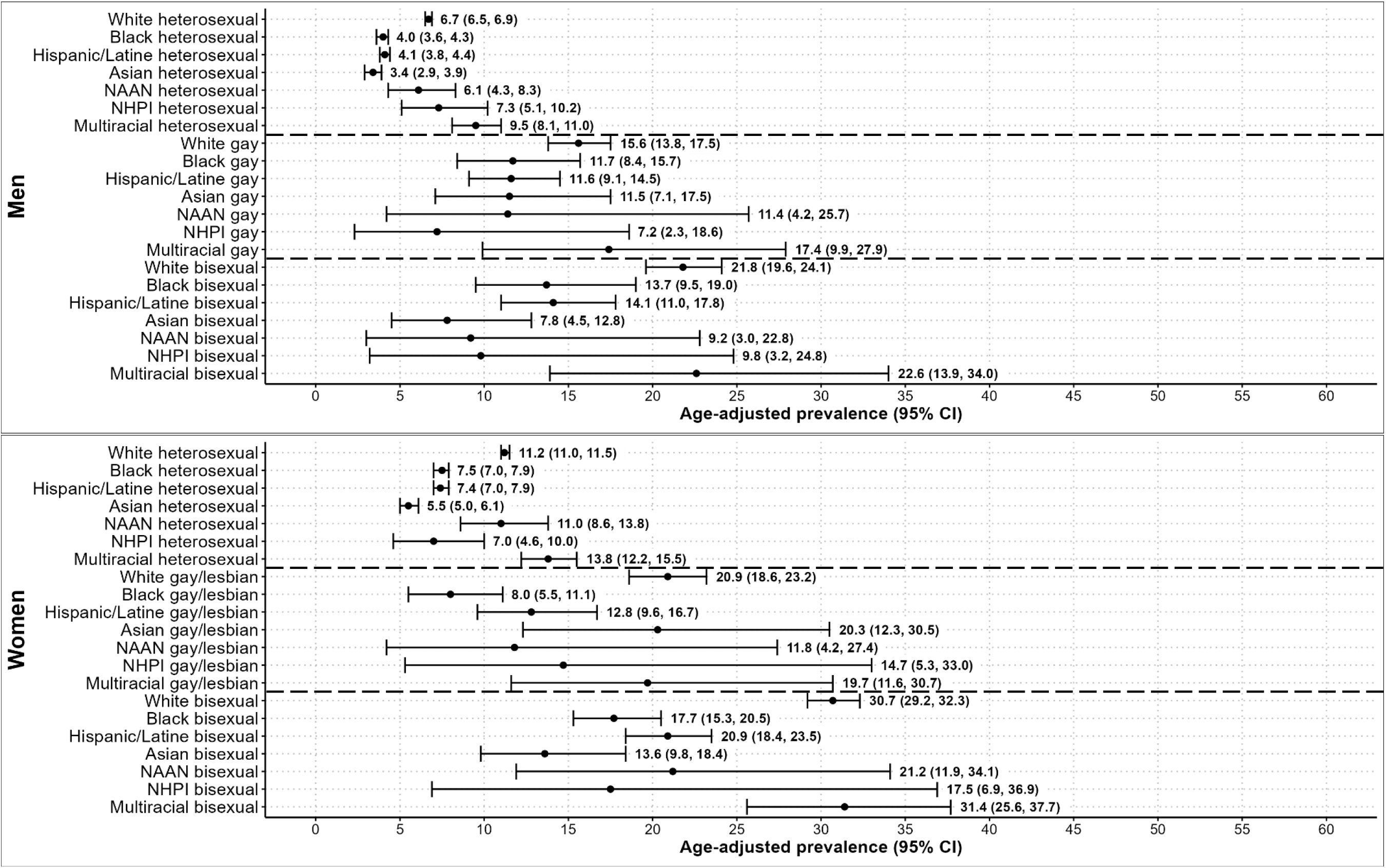
Age-adjusted prevalence of past-year major depressive episode among US adults, NSDUH 2015-2020 (n=234,374) *Note:* NSDUH = National Survey on Drug Use and Health. CI = credible interval. NAAN = Native American or American Indian. NHPI = Native Hawaiian or Pacific Islander. Estimates are design-weighted to account for NSDUH complex sample survey weights.

**Figure 2.**
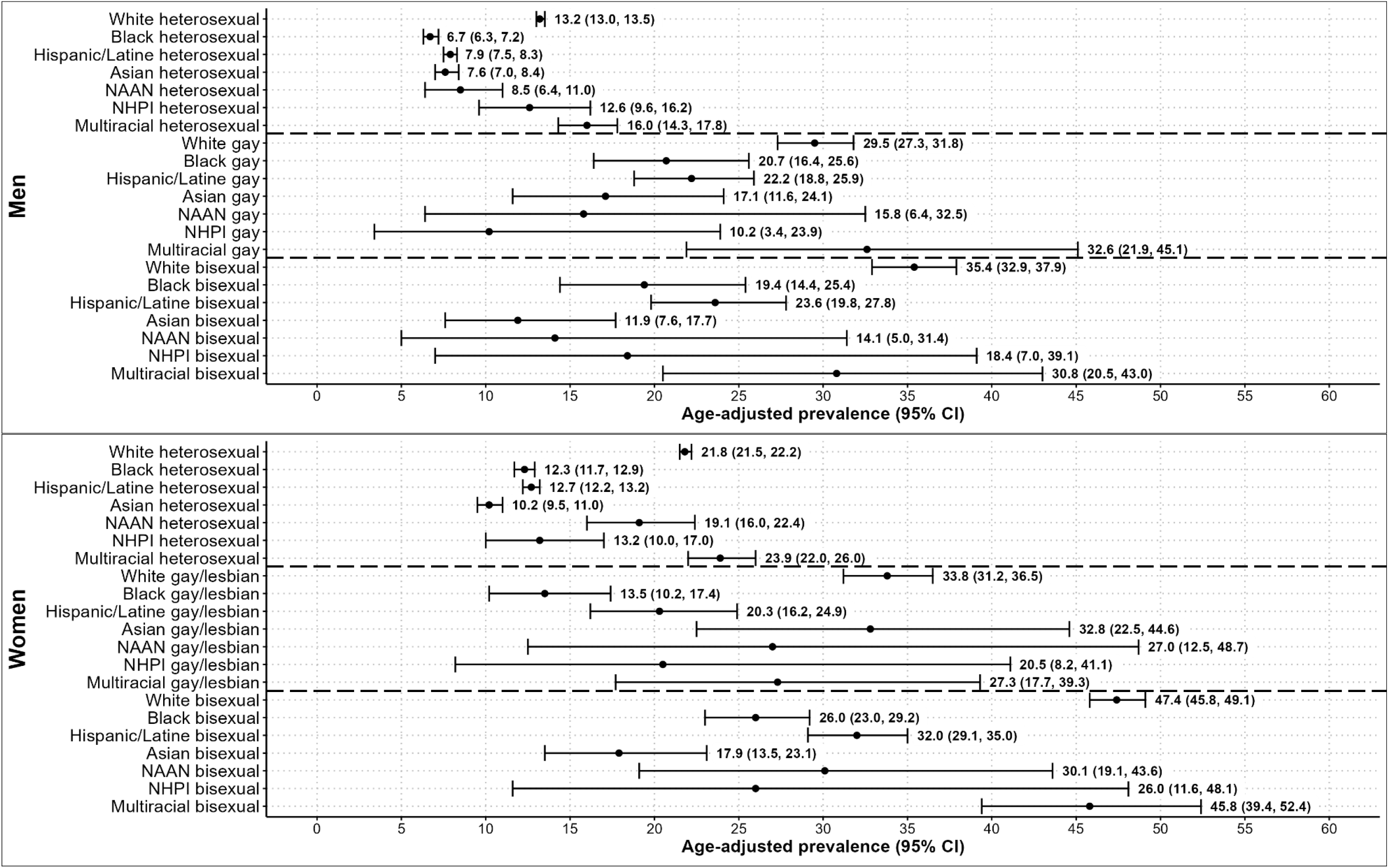
Age-adjusted prevalence of lifetime major depressive episode among US adults, NSDUH 2015-2020 (n=234,722) *Note*: NSDUH = National Survey on Drug Use and Health. CI = credible interval. NAAN = Native American or American Indian. NHPI = Native Hawaiian or Pacific Islander. Estimates are design-weighted to account for NSDUH complex sample survey weights.

In unidimensional (i.e., non-intersectional) terms and relative to White people, Asian people (OR=0.46, 95% CI: 0.38–0.56), Black people (OR=0.55, 95% CI: 0.46–0.65), Hispanic/Latine people (OR=0.61, 95% CI: 0.52–0.71), and Native Hawaiian or Pacific Islander people (OR=0.71, 95% CI: 0.50–0.98) had lower odds of past-year MDE (Model 3 in Table 2), while Multiracial people (OR=1.26, 95% CI: 1.04–1.52) had increased odds of past-year MDE. Estimates were similar for lifetime MDE, except Multiracial people (OR=1.08, 95% CI: 0.79–1.45) had similar odds and Native American or Alaska Native people (OR=0.58, 95% CI: 0.38–0.86) had lower odds compared to White people (Model 3 in Table 3). For past-year and lifetime MDE, women (ORs=1.51–1.60), gay/lesbian people (ORs=2.19–2.27), and bisexual people (ORs=2.78–3.33) had elevated odds.

Sample-level intersectionality measures showed that race/ethnicity, sex/gender, and sexual orientation jointly structure the social patterning of MDE among US adults. Model 1 VPCs indicated that 12.8% (past-year) and 12.6% (lifetime) of the variance in MDE is attributable to differences across the intersectional groups. Relative to intermediary models adjusting for race/ethnicity (Model 2a PCVs = 10.0–17.1%) and sex/gender (Model 2b PCVs = 7.5–7.9%), sexual orientation (Model 2c PCVs = 42.9–54.0%) explained a higher proportion of the Model 1 between-group variance in MDE prevalence. The intersectional interaction model (Model 3) indicated that most, but not all, of the intersectional group-level variance in past-year (PCV = 96.8%) and lifetime (PCV = 87.9%) MDE were explained by the identity variable additive effects. The remaining differences between intersectional groups (i.e., 100% minus the PCV) in the prevalence of past-year (3.2%) and lifetime (12.1%) MDE was attributable to intersectional effects.

Table 4 shows group-specific estimates of excess/reduced prevalence. In general, estimates for lifetime MDE had a larger absolute value compared to past-year MDE, and this pattern is supported by greater group-level variance (lifetime = 0.05; past-year = 0.01) that remained unexplained in the fully-adjusted model (Model 3 in Tables 2 and 3). For lifetime MDE, a significant intersectional effect was present for Black gay/lesbian women whereby the model-predicted prevalence was -4.6 (95% CI: -9.4%, -0.7%) percentage points lower than expected prevalence based on identity variable additive effects. Although no other estimates were significant at a canonical 95% CI level, we note intersectional groups with highest excess prevalence of lifetime MDE (i.e., higher than expected prevalence) were Asian gay/lesbian women (4.5%, 95% CI: -1.0%, 11.8%), White bisexual women (3.9%, 95% CI: -1.9%, 9.6%), and Black gay men (3.1%, 95% CI: -0.4%, 7.1%). Conversely, groups with greatest reduced prevalence of lifetime MDE (i.e., lower than expected prevalence) were Black gay/lesbian women, Asian bisexual women (−3.5%, 95% CI: -8.7%, 1.1%), and Multiracial gay/lesbian women (−3.3%, 95% CI: -11.6%, 4.4%).

**Table 4.** Age-adjusted excess/reduced prevalence due to interaction of race/ethnicity, sex/gender, and sexual orientation.

| Intersectional group | Past-year MDE |  | Lifetime MDE |  |
| --- | --- | --- | --- | --- |
|  | MP (95% CI) | ERP (95% CI) | MP (95% CI) | ERP (95% CI) |
| White heterosexual men | 6.7 (6.5, 6.9) | -0.4 (-1.1, 0.2) | 13.2 (13.0, 13.5) | -1.8 (-4.3, 0.5) |
| White gay men | 15.6 (13.8, 17.5) | 0.2 (-1.4, 1.8) | 29.5 (27.3, 31.8) | 0.9 (-3.7, 5.5) |
| White bisexual men | 21.8 (19.6, 24.1) | 0.6 (-1.4, 2.8) | 35.4 (32.9, 37.9) | 1.6 (-3.6, 7.0) |
| White heterosexual women | 11.2 (11.0, 11.5) | 0.0 (-1.0, 1.0) | 21.8 (21.5, 22.2) | 0.2 (-3.5, 3.4) |
| White gay/lesbian women | 20.9 (18.6, 23.2) | -0.6 (-2.9, 1.5) | 33.8 (31.2, 36.5) | -3.1 (-9.0, 2.3) |
| White bisexual women | 30.7 (29.2, 32.3) | 1.0 (-1.5, 3.6) | 47.4 (45.8, 49.1) | 3.9 (-1.9, 9.6) |
| Black heterosexual men | 4.0 (3.6, 4.3) | -0.1 (-0.6, 0.3) | 6.7 (6.3, 7.2) | -0.6 (-2.1, 0.6) |
| Black gay men | 11.7 (8.4, 15.7) | 0.5 (-0.7, 2.0) | 20.7 (16.4, 25.6) | 3.1 (-0.4, 7.1) |
| Black bisexual men | 13.7 (9.5, 19.0) | 0.2 (-1.4, 2.1) | 19.4 (14.4, 25.4) | 0.6 (-3.5, 4.9) |
| Black heterosexual women | 7.5 (7.0, 7.9) | 0.6 (-0.1, 1.3) | 12.3 (11.7, 12.9) | 1.1 (-1.1, 2.8) |
| Black gay/lesbian women | 8.0 (5.5, 11.1) | -1.3 (-3.4, 0.3) | 13.5 (10.2, 17.4) | **-4.6 (-9.4, -0.7)** |
| Black bisexual women | 17.7 (15.3, 20.5) | -0.3 (-2.3, 1.7) | 26.0 (23.0, 29.2) | 0.5 (-4.2, 4.9) |
| Hispanic/Latine heterosexual men | 4.1 (3.8, 4.4) | -0.3 (-0.8, 0.1) | 7.9 (7.5, 8.3) | -1.1 (-2.9, 0.5) |
| Hispanic/Latine gay men | 11.6 (9.1, 14.5) | 0.4 (-0.7, 1.9) | 22.2 (18.8, 25.9) | 2.8 (-0.9, 6.6) |
| Hispanic/Latine bisexual men | 14.1 (11.0, 17.8) | 0.1 (-1.5, 1.9) | 23.6 (19.8, 27.8) | 1.2 (-3.1, 5.6) |
| Hispanic/Latine heterosexual women | 7.4 (7.0, 7.9) | 0.2 (-0.5, 0.9) | 12.7 (12.2, 13.2) | -0.5 (-3.3, 1.6) |
| Hispanic/Latine gay/lesbian women | 12.8 (9.6, 16.7) | -0.4 (-2.3, 1.3) | 20.3 (16.2, 24.9) | -2.9 (-8.1, 1.5) |
| Hispanic/Latine bisexual women | 20.9 (18.4, 23.5) | 0.4 (-1.6, 2.5) | 32.0 (29.1, 35.0) | 1.8 (-3.2, 6.6) |
| Asian heterosexual men | 3.4 (2.9, 3.9) | -0.1 (-0.5, 0.3) | 7.6 (7.0, 8.4) | 0.4 (-1.1, 1.7) |
| Asian gay men | 11.5 (7.1, 17.5) | 0.3 (-0.7, 1.7) | 17.1 (11.6, 24.1) | 1.0 (-2.5, 5.1) |
| Asian bisexual men | 7.8 (4.5, 12.8) | -0.3 (-2.0, 1.1) | 11.9 (7.6, 17.7) | -2.4 (-6.5, 1.4) |
| Asian heterosexual women | 5.5 (5.0, 6.1) | 0.0 (-0.7, 0.6) | 10.2 (9.5, 11.0) | -0.1 (-2.4, 1.7) |
| Asian gay/lesbian women | 20.3 (12.3, 30.5) | 0.7 (-0.9, 3.2) | 32.8 (22.5, 44.6) | 4.5 (-1.0, 11.8) |
| Asian bisexual women | 13.6 (9.8, 18.4) | -0.5 (-2.6, 1.4) | 17.9 (13.5, 23.1) | -3.5 (-8.7, 1.1) |
| NAAN heterosexual men | 6.1 (4.3, 8.3) | -0.1 (-1.0, 0.7) | 8.5 (6.4, 11.0) | -1.2 (-3.8, 1.1) |
| NAAN gay men | 11.4 (4.2, 25.7) | 0.0 (-1.9, 2.1) | 15.8 (6.4, 32.5) | -0.5 (-5.8, 5.6) |
| NAAN bisexual men | 9.2 (3.0, 22.8) | -0.2 (-2.8, 2.5) | 14.1 (5.0, 31.4) | -1.1 (-7.5, 5.8) |
| NAAN heterosexual women | 11.0 (8.6, 13.8) | 0.4 (-0.9, 1.8) | 19.1 (16.0, 22.4) | 2.7 (-0.9, 6.3) |
| NAAN gay/lesbian women | 11.8 (4.2, 27.4) | -0.1 (-2.8, 2.9) | 27.0 (12.5, 48.7) | 0.9 (-6.2, 9.3) |
| NAAN bisexual women | 21.2 (11.9, 34.1) | 0.0 (-3.4, 3.6) | 30.1 (19.1, 43.6) | 0.4 (-6.9, 8.4) |
| NHPI heterosexual men | 7.3 (5.1, 10.2) | 0.2 (-0.5, 1.2) | 12.6 (9.6, 16.2) | 1.4 (-1.2, 4.2) |
| NHPI gay men | 7.2 (2.3, 18.6) | -0.1 (-1.8, 1.6) | 10.2 (3.4, 23.9) | -1.4 (-6.5, 4.0) |
| NHPI bisexual men | 9.8 (3.2, 24.8) | -0.1 (-2.3, 2.3) | 18.4 (7.0, 39.1) | -0.1 (-6.1, 6.8) |
| NHPI heterosexual women | 7.0 (4.6, 10.0) | -0.2 (-1.5, 0.8) | 13.2 (10.0, 17.0) | -0.5 (-4.1, 2.8) |
| NHPI gay/lesbian women | 14.7 (5.3, 33.0) | 0.1 (-2.3, 2.8) | 20.5 (8.2, 41.1) | -0.1 (-6.8, 7.6) |
| NHPI bisexual women | 17.5 (6.9, 36.9) | 0.1 (-2.9, 3.7) | 26.0 (11.6, 48.1) | 0.5 (-7.1, 9.2) |
| Multiracial heterosexual men | 9.5 (8.1, 11.0) | 0.2 (-0.9, 1.2) | 16.0 (14.3, 17.8) | -0.3 (-3.7, 2.7) |
| Multiracial gay men | 17.4 (9.9, 27.9) | 0.1 (-2.3, 2.9) | 32.6 (21.9, 45.1) | 1.8 (-4.9, 9.6) |
| Multiracial bisexual men | 22.6 (13.9, 34.0) | 0.2 (-2.9, 3.6) | 30.8 (20.5, 43.0) | -0.6 (-8.0, 7.1) |
| Multiracial heterosexual women | 13.8 (12.2, 15.5) | 0.1 (-1.4, 1.6) | 23.9 (22.0, 26.0) | 0.9 (-3.5, 4.7) |
| Multiracial gay/lesbian women | 19.7 (11.6, 30.7) | -0.3 (-3.8, 3.1) | 27.3 (17.7, 39.3) | -3.3 (-11.6, 4.4) |
| Multiracial bisexual women | 31.4 (25.6, 37.7) | -0.5 (-4.2, 3.2) | 45.8 (39.4, 52.4) | 1.4 (-5.8, 8.8) |

Note: MDE = Major depressive episode. MP = Model-predicted prevalence. ERP = Excess or reduced prevalence due to interaction. CI = Credible interval. NAAN = Native American or Alaska Native. NHPI = Native Hawaiian or Pacific Islander. ERP values represent percentage point increases/decreases. Positive ERP values (shaded) indicate excess prevalence (i.e., greater than expected prevalence), while negative ERP values (unshaded) indicate reduced prevalence (i.e., lower than expected prevalence).

## DISCUSSION

The current study examined how race/ethnicity, sex/gender, and sexual orientation intersect to structure population-level patterns of past-year and lifetime MDE. Using design-weighted intersectional MAIDHA with a nationally representative sample of US adults, significant main effects indicated that people who were Multiracial, White, women, gay/lesbian, or bisexual had greater odds of MDE, which is broadly consistent with prior documentation of inequities by race/ethnicity,^7–14^ sex/gender,^8,15–18^ and sexual orientation.^19–21^ We extend prior work by characterizing a complex patterning with prevalence estimates ranging from 3.4% (Asian heterosexual men) to 31.4% (Multiracial bisexual women) for past-year MDE and from 6.7% (Black heterosexual men) to 47.4% (White bisexual women) for lifetime MDE. Our results highlight how non-intersectional methods may conceal within- and between-group heterogeneity. For example, comparing racial/ethnic and sex/gender groups within overall sexual orientation categories, we found highly variable prevalence estimates among heterosexual (past-year: 3.4–13.8%; lifetime: 6.7–23.9%), gay/lesbian (past-year: 7.2–20.9%; lifetime: 10.2–33.8%), and bisexual (past-year: 7.8–31.4%; lifetime: 11.9–47.4%) adults.

We quantified the amount of between-group variance in depression prevalence explained by additive effects (i.e., the main effects of race/ethnicity + sex/gender + sexual orientation) and by intersectional effects (i.e., excess/reduced prevalence due to interaction of the constituent identity variables). Fully adjusted model results showed that additive effects explained the majority of between-group variance, while approximately 3% (past-year) and 12% (lifetime) of differences were attributable to intersectional effects. When assessing the relative roles of the identity variables, we found that models adjusting only for sexual orientation explained more between-group variance compared to models adjusting for only race/ethnicity or only sex/gender. These findings suggest that sexual orientation plays a pronounced role driving US adult depression prevalence inequities. This was further evidenced by relatively large main effect estimates for gay/lesbian and bisexual identity and elevated prevalence estimates for intersectional groups inclusive of gay/lesbian and bisexual people.

As posited by minority stress theory^46,47^ and the ecosocial theory of disease distribution,^48^ heterogeneous depression prevalence may be due to inequitable distribution of power and resources (e.g., social/economic deprivation, discrimination, exposure to hazardous environments). Resultantly, structurally marginalized and minoritized groups may have increased depression prevalence arising from long-term adverse exposures leading to increased stress, maladaptive coping, and other proximal depression risk factors. Our findings regarding intersectional group-specific excess/reduced prevalence further illustrate this heterogeneity. In general, for both past-year and lifetime MDE across intersectional groups, bisexual women and gay men had excess MDE prevalence and gay/lesbian women had reduced MDE prevalence.

From an intersectional perspective,^27,28^ the complex patterning observed in the current study may be a function of each group’s unique exposures and experiences as well as its position within US social hierarchies. For example, certain groups, such as Black gay/lesbian women, had lower than expected prevalence of lifetime MDE. This may indicate that belonging in these groups may come with certain protective qualities (e.g., social support, reduced exposure to stressors); however, we note that for many such groups, prevalence estimates were still higher than the population average. Our results also share commonalities with a prior study examining lifetime suicidality at the intersection of race/ethnicity, sex/gender, and sexual orientation.^49^ Both the current and prior study found that, relative to White heterosexual men, both Black and Hispanic/Latine heterosexual men had lower prevalence of mental health concerns, but racial/ethnic differences were attenuated among sexual minority men. Therefore, risk factors for adverse mental health outcomes may be compounded in certain intersectional groups (e.g., Hispanic/Latine sexual minority men).

Taken together, our findings highlight that depression prevention, screening, and treatment efforts among sexual minority people are particularly warranted. Identifying and addressing structural mechanisms (e.g., heterosexist laws/policies) that may drive these inequities is critical to reduce depression prevalence and incidence. In addition, healthcare systems should routinize collection of sexual orientation and gender identity (SOGI) data in clinical settings and ensure clinician proficiency to provide identity-affirming mental health services.^50,51^ To this aim, SAMHSA has compiled relevant training curricula for LGBTQ+ affirming primary and behavioral healthcare.^52^ Prior work has shown that sexual minority adults face many of the same barriers as heterosexual adults when seeking mental health services, including affordability and availability,^53^ but may also experience unique treatment barriers, such as sexual identity disclosure and finding an affirming provider.^54^ Overall, continued policy efforts are needed to reduce intersectional inequities in depression and ensure that those with depression can access treatment.

### Limitations & Strengths

Findings should be considered in light of certain limitations. First, we were limited to broad racial/ethnic categories which may conceal meaningful within-group heterogeneity. Second, NSDUH interviewers assigned participants’ sex/gender (limited to “male” and “female” responses) presumably based on social cues (e.g., vocal timbre, gender expression). While this method is common in survey research,^23^ it may introduce measurement error by treating sex and gender as a unidimensional construct, introducing misclassification bias (e.g., nonbinary participants misclassified as male or female), and neglecting sex/gender diversity (e.g., no options for intersex and transgender people). Third, for sexual orientation, participants had limited response options, so those with unlisted identities (e.g., queer, asexual) may have refused response or selected an option unreflective of their identity. Finally, due to smaller sample sizes, intersectional groups with lesbian, gay, and bisexual people had reduced statistical precision.

The current study also provides notable innovations. While previous applications have used complex sample survey data,^34,55^ this is the first to conduct a design-weighted MAIDHA with Bayesian statistical methods. Our approach opens opportunities for future researchers to efficiently estimate intersectional effects for health outcomes using representative datasets, including NSDUH and other federal health surveys. Moreover, Bayesian analysis has flexible estimation procedures with small subgroup sample sizes commonly found in quantitative intersectional research. Additional strengths include using racial/ethnic (i.e., Native Hawaiian and Pacific Islander, Native American and Alaska Native) and sexual orientation (i.e., gay/lesbian and bisexual) categories often excluded or combined into catch-all groups (e.g., sexual minority, “other” race) to provide prevalence estimates for understudied population subgroups (e.g., Native American and Alaska Native bisexual women).

## Conclusions

Our findings document substantial inequities in the prevalence of past-year and lifetime MDE among US adults. We provide strong evidence that examining social identity groups in isolation conceals within-group heterogeneity (e.g., sexual orientation and sex/gender differences within racial/ethnic groups). Additive effects accounted for most intersectional group prevalence differences; however, intersectional effects were present whereby some groups, particularly for lifetime MDE, had evidence of excess/reduced prevalence beyond those estimated by additive effects alone. Sexual orientation, relative to race/ethnicity and sex/gender, emerged as a stronger determinant of intersectional depression inequities, which may highlight the outsized role of sexual orientation-related structural factors (e.g., sexual orientation-based discrimination, heterosexist laws/policies). Our study highlights the promise of using quantitative intersectional methods to identify disproportionately burdened population subgroups and inform intervention and prevention efforts to reduce depression-related costs, morbidity, and mortality. Looking ahead, we encourage future researchers and policy actors to identify and intervene upon structural mechanisms driving these inequities.

## Data Availability

All data and R code are available online at: https://github.com/fhmcguire/NSDUH_intersectional_depression

## References

1. Varghese S, Frey BN, Schneider MA, Kapczinski F, de Azevedo Cardoso T. Functional and cognitive impairment in the first episode of depression: A systematic review. Acta Psychiatr Scand. 2022;145(2):156–185. doi:10.1111/acps.13385

2. Hunt GE, Malhi GS, Lai HMX, Cleary M. Prevalence of comorbid substance use in major depressive disorder in community and clinical settings, 1990-2019: Systematic review and meta-analysis. J Affect Disord. 2020;266:288–304. doi:10.1016/j.jad.2020.01.141

3. Cuijpers P, Vogelzangs N, Twisk J, Kleiboer A, Li J, Penninx BW. Comprehensive Meta-Analysis of Excess Mortality in Depression in the General Community Versus Patients With Specific Illnesses. Am J Psychiatry. 2014;171(4):453–462. doi:10.1176/appi.ajp.2013.13030325

4. Moitra M, Santomauro D, Degenhardt L, et al. Estimating the risk of suicide associated with mental disorders: A systematic review and meta-regression analysis. J Psychiatr Res. 2021;137:242–249. doi:10.1016/j.jpsychires.2021.02.053

5. Greenberg PE, Fournier AA, Sisitsky T, et al. The Economic Burden of Adults with Major Depressive Disorder in the United States (2010 and 2018). PharmacoEconomics. 2021;39(6):653–665. doi:10.1007/s40273-021-01019-4

6. Substance Abuse and Mental Health Services Administration. Key Substance Use and Mental Health Indicators in the United States: Results from the 2020 National Survey on Drug Use and Health (HHS Publication No. PEP21-07-01-003, NSDUH Series H-56). Center for Behavioral Health Statistics and Quality, Substance Abuse and Mental Health Services Administration; 2021. https://www.samhsa.gov/data/

7. Kessler RC, Berglund P, Demler O, et al. The epidemiology of major depressive disorder: results from the National Comorbidity Survey Replication (NCS-R). JAMA. 2003;289(23):3095–3105. doi:10.1001/jama.289.23.3095

8. Hasin DS, Sarvet AL, Meyers JL, et al. Epidemiology of Adult DSM-5 Major Depressive Disorder and Its Specifiers in the United States. JAMA Psychiatry. 2018;75(4):336–346. doi:10.1001/jamapsychiatry.2017.4602

9. Riolo SA, Nguyen TA, Greden JF, King CA. Prevalence of depression by race/ethnicity: findings from the National Health and Nutrition Examination Survey III. Am J Public Health. 2005;95(6):998–1000. doi:10.2105/AJPH.2004.047225

10. Hooker K, Phibbs S, Irvin VL, et al. Depression Among Older Adults in the United States by Disaggregated Race and Ethnicity. The Gerontologist. 2019;59(5):886–891. doi:10.1093/geront/gny159

11. González HM, Tarraf W, Whitfield KE, Vega WA. The epidemiology of major depression and ethnicity in the United States. J Psychiatr Res. 2010;44(15):1043–1051. doi:10.1016/j.jpsychires.2010.03.017

12. Blazer DG, Kessler RC, McGonagle KA, Swartz MS. The prevalence and distribution of major depression in a national community sample: the National Comorbidity Survey. Am J Psychiatry. 1994;151(7):979–986. doi:10.1176/ajp.151.7.979

13. Williams DR, González HM, Neighbors H, et al. Prevalence and distribution of major depressive disorder in African Americans, Caribbean blacks, and non-Hispanic whites: results from the National Survey of American Life. Arch Gen Psychiatry. 2007;64(3):305–315. doi:10.1001/archpsyc.64.3.305

14. Breslau J, Kendler KS, Su M, Gaxiola-Aguilar S, Kessler RC. Lifetime risk and persistence of psychiatric disorders across ethnic groups in the United States. Psychol Med. 2005;35(3):317–327. doi:10.1017/s0033291704003514

15. Kessler RC, McGonagle KA, Swartz M, Blazer DG, Nelson CB. Sex and depression in the National Comorbidity Survey. I: Lifetime prevalence, chronicity and recurrence. J Affect Disord. 1993;29(2-3):85–96. doi:10.1016/0165-0327(93)90026-g

16. Kessler RC. Epidemiology of women and depression. J Affect Disord. 2003;74(1):5–13. doi:10.1016/s0165-0327(02)00426-3

17. Inaba A, Thoits PA, Ueno K, Gove WR, Evenson RJ, Sloan M. Depression in the United States and Japan: gender, marital status, and SES patterns. Soc Sci Med 1982. 2005;61(11):2280–2292. doi:10.1016/j.socscimed.2005.07.014

18. Salk RH, Hyde JS, Abramson LY. Gender differences in depression in representative national samples: Meta-analyses of diagnoses and symptoms. Psychol Bull. 2017;143(8):783–822. doi:10.1037/bul0000102

19. Li G, Pollitt AM, Russell ST. Depression and Sexual Orientation During Young Adulthood: Diversity Among Sexual Minority Subgroups and the Role of Gender Nonconformity. Arch Sex Behav. 2016;45(3):697–711. doi:10.1007/s10508-015-0515-3

20. Chaudhry AB, Reisner SL. Disparities by Sexual Orientation Persist for Major Depressive Episode and Substance Abuse or Dependence: Findings from a National Probability Study of Adults in the United States. LGBT Health. 2019;6(5):261–266. doi:10.1089/lgbt.2018.0207

21. Scott RL, Lasiuk G, Norris C. The relationship between sexual orientation and depression in a national population sample. J Clin Nurs. 2016;25(23-24):3522–3532. doi:10.1111/jocn.13286

22. Bostwick WB, Boyd CJ, Hughes TL, McCabe SE. Dimensions of sexual orientation and the prevalence of mood and anxiety disorders in the United States. Am J Public Health. 2010;100(3):468–475. doi:10.2105/AJPH.2008.152942

23. Bauer GR. Sex and Gender Multidimensionality in Epidemiologic Research. Am J Epidemiol. 2023;192(1):122–132. doi:10.1093/aje/kwac173

24. Homan P. Structural Sexism and Health in the United States: A New Perspective on Health Inequality and the Gender System. Am Sociol Rev. 2019;84(3):486–516. doi:10.1177/0003122419848723

25. Bailey ZD, Krieger N, Agénor M, Graves J, Linos N, Bassett MT. Structural racism and health inequities in the USA: evidence and interventions. Lancet Lond Engl. 2017;389(10077):1453–1463. doi:10.1016/S0140-6736(17)30569-X

26. Hatzenbuehler ML. Structural Stigma and Health Inequalities: Research Evidence and Implications for Psychological Science. Am Psychol. 2016;71(8):742–751. doi:10.1037/amp0000068

27. Crenshaw K. Demarginalizing the Intersection of Race and Sex: A Black Feminist Critique of Antidiscrimination Doctrine, Feminist Theory and Antiracist Politics. Univ Chic Leg Forum. 1989;1989(1).

28. Crenshaw K. Mapping the Margins: Intersectionality, Identity Politics, and Violence against Women of Color. Stanford Law Rev. 1991;43(6):1241–1299. doi:10.2307/1229039

29. Bowleg L. The Problem With the Phrase Women and Minorities: Intersectionality—an Important Theoretical Framework for Public Health. Am J Public Health. 2012;102(7):1267–1273. doi:10.2105/AJPH.2012.300750

30. Bowleg L. When Black + Lesbian + Woman ≠ Black Lesbian Woman: The Methodological Challenges of Qualitative and Quantitative Intersectionality Research. Sex Roles. 2008;59(5):312–325. doi:10.1007/s11199-008-9400-z

31. Bauer GR, Churchill SM, Mahendran M, Walwyn C, Lizotte D, Villa-Rueda AA. Intersectionality in quantitative research: A systematic review of its emergence and applications of theory and methods. SSM - Popul Health. 2021;14:100798. doi:10.1016/j.ssmph.2021.100798

32. Rodriguez-Seijas C, Eaton NR, Pachankis JE. Prevalence of psychiatric disorders at the intersection of race and sexual orientation: Results from the National Epidemiologic Survey of Alcohol and Related Conditions-III. J Consult Clin Psychol. 2019;87(4):321–331. doi:10.1037/ccp0000377

33. Evans CR, Williams DR, Onnela JP, Subramanian SV. A multilevel approach to modeling health inequalities at the intersection of multiple social identities. Soc Sci Med. 2018;203:64–73. doi:10.1016/j.socscimed.2017.11.011

34. Evans CR, Erickson N. Intersectionality and depression in adolescence and early adulthood: A MAIHDA analysis of the national longitudinal study of adolescent to adult health, 1995-2008. Soc Sci Med 1982. 2019;220:1–11. doi:10.1016/j.socscimed.2018.10.019

35. Merlo J. Multilevel analysis of individual heterogeneity and discriminatory accuracy (MAIHDA) within an intersectional framework. Soc Sci Med 1982. 2018;203:74–80. doi:10.1016/j.socscimed.2017.12.026

36. Center for Behavioral Health Statistics and Quality. 2019 National Survey on Drug Use and Health (NSDUH): Methodological Summary and Definitions. Substance Abuse and Mental Health Services Administration; 2020. Accessed December 16, 2021. https://www.samhsa.gov/data/

37. Substance Abuse and Mental Health Services Administration. National Survey on Drug Use and Health (NSDUH). Published 2022. Accessed May 24, 2022. https://www.samhsa.gov/data/data-we-collect/nsduh-national-survey-drug-use-and-health

38. American Psychiatric Association. Diagnostic and Statistical Manual of Mental Disorders. 5th ed.; 2013. https://doi.org/10.1176/appi.books.9780890425596

39. Jones JM. LGBT Identification in U.S. Ticks Up to 7.1%. Gallup.com. Published February 17, 2022. Accessed February 14, 2023. https://news.gallup.com/poll/389792/lgbt-identification-ticks-up.aspx

40. Frey WH. Less than half of US children under 15 are white, census shows. Brookings. Published June 24, 2019. Accessed April 13, 2023. https://www.brookings.edu/research/less-than-half-of-us-children-under-15-are-white-census-shows/

41. Hespanhol L, Vallio CS, Costa LM, Saragiotto BT. Understanding and interpreting confidence and credible intervals around effect estimates. Braz J Phys Ther. 2019;23(4):290–301. doi:10.1016/j.bjpt.2018.12.006

42. McElreath R. Statistical Rethinking: A Bayesian Course with Examples in R and Stan. 2nd ed. CRC Press; 2020.

43. Williams MR, Savitsky TD. Bayesian Estimation Under Informative Sampling with Unattenuated Dependence. Bayesian Anal. 2020;15(1):57–77. doi:10.1214/18-BA1143

44. Lumley T. Package “survey.” Published online 2021. Accessed January 14, 2022. https://cran.r-project.org/web/packages/survey/survey.pdf

45. Williams MR. csSampling: Complex survey sampling. Published online 2022. Accessed January 6, 2023. https://rdrr.io/github/RyanHornby/csSampling/

46. Brooks V. Minority Stress and Lesbian Women. Lexington Books; 1981.

47. Meyer IH. Prejudice, Social Stress, and Mental Health in Lesbian, Gay, and Bisexual Populations: Conceptual Issues and Research Evidence. Psychol Bull. 2003;129(5):674–697. doi:10.1037/0033-2909.129.5.674

48. Krieger N. Theories for social epidemiology in the 21st century: an ecosocial perspective. Int J Epidemiol. 2001;30(4):668–677. doi:10.1093/ije/30.4.668

49. Ramchand R, Schuler MS, Schoenbaum M, Colpe L, Ayer L. Suicidality Among Sexual Minority Adults: Gender, Age, and Race/Ethnicity Differences. Am J Prev Med. 2022;62(2):193–202. doi:10.1016/j.amepre.2021.07.012

50. Cahill S, Makadon H. Sexual Orientation and Gender Identity Data Collection in Clinical Settings and in Electronic Health Records: A Key to Ending LGBT Health Disparities. LGBT Health. 2014;1(1):34–41. doi:10.1089/lgbt.2013.0001

51. Cahill S, Singal R, Grasso C, et al. Do ask, do tell: high levels of acceptability by patients of routine collection of sexual orientation and gender identity data in four diverse American community health centers. PloS One. 2014;9(9):e107104. doi:10.1371/journal.pone.0107104

52. Substance Abuse and Mental Health Services Administration. LGBT Training Curricula for Behavioral Health and Primary Care Practitioners. Published May 27, 2014. Accessed April 7, 2023. https://www.samhsa.gov/behavioral-health-equity/lgbtqi/curricula

53. Coombs NC, Meriwether WE, Caringi J, Newcomer SR. Barriers to healthcare access among U.S. adults with mental health challenges: A population-based study. SSM - Popul Health. 2021;15:100847. doi:10.1016/j.ssmph.2021.100847

54. Romanelli M, Hudson KD. Individual and systemic barriers to health care: Perspectives of lesbian, gay, bisexual, and transgender adults. Am J Orthopsychiatry. 2017;87:714–728. doi:10.1037/ort0000306

55. Persmark A, Wemrell M, Evans CR, Subramanian SV, Leckie G, Merlo J. Intersectional inequalities and the U.S. opioid crisis: challenging dominant narratives and revealing heterogeneities. Crit Public Health. 2020;30(4):398–414. doi:10.1080/09581596.2019.1626002

