## Supplementary Materials for "Depression at the intersection of race/ethnicity, sex/gender, and sexual orientation in a nationally representative sample of US adults: A design-weighted MAIHDA"

### Table of Contents

Model equations ........ 2

Table S1: Intersectional group frequencies ........ 3

### Model equations

Let *i* = individuals and *j* = intersectional group:

**Model 1: Age-adjusted prevalence model**

$$Logit(Depression_{ij}) = (\gamma_{00} + \gamma_{01}Age2634_i + \gamma_{02}Age3549_i + \gamma_{03}Age50plus_i) + (u_{0j} + r_{ij})$$

**Model 2a: Race/ethnicity and age-adjusted model**

$$Logit(Depression_{ij}) = (\gamma_{00} + \gamma_{01}Age2634_i + \gamma_{02}Age3549_i + \gamma_{03}Age50plus_i + \gamma_{04}Asian_i + \gamma_{05}Black_i + \gamma_{06}NativeAmerican_i + \gamma_{07}NativeHawaiianPI_i + \gamma_{08}Multiracial_i + \gamma_{09}Latine_i) + (u_{0j} + r_{ij})$$

**Model 2b: Sex/Gender and age-adjusted model**

$$Logit(Depression_{ij}) = (\gamma_{00} + \gamma_{01}Age2634_i + \gamma_{02}Age3549_i + \gamma_{03}Age50plus_i + \gamma_{04}Woman_i) + (u_{0j} + r_{ij})$$

**Model 2c: Sexual orientation and age-adjusted model**

$$Logit(Depression_{ij}) = (\gamma_{00} + \gamma_{01}Age2634_i + \gamma_{02}Age3549_i + \gamma_{03}Age50plus_i + \gamma_{04}GayLesbian_i + \gamma_{05}Bisexual_i) + (u_{0j} + r_{ij})$$

**Model 3: Fully adjusted model (intersectional interaction)**

$$Logit(Depression_{ij}) = (\gamma_{00} + \gamma_{01}Age2634_i + \gamma_{02}Age3549_i + \gamma_{03}Age50plus_i + \gamma_{04}Asian_i + \gamma_{05}Black_i + \gamma_{06}NativeAmerican_i + \gamma_{07}NativeHawaiianPI_i + \gamma_{08}Multiracial_i + \gamma_{09}Latine_i + \gamma_{09}Woman_i + \gamma_{10}GayLesbian_i + \gamma_{11}Bisexual_i) + (u_{0j} + r_{ij})$$

### Table S1: Intersectional group frequencies

| ID | Intersectional Group | Lifetime MDE (n=234,722) | Past-year MDE (n=234,374) |
|---|---|---|---|
| 1 | White heterosexual men | 64392 | 64331 |
| 2 | White gay men | 1584 | 1581 |
| 3 | White bisexual men | 1679 | 1675 |
| 4 | White heterosexual women | 69450 | 69329 |
| 5 | White gay/lesbian women | 1459 | 1458 |
| 6 | White bisexual women | 5656 | 5630 |
| 7 | Black heterosexual men | 11914 | 11905 |
| 8 | Black gay men | 323 | 323 |
| 9 | Black bisexual men | 248 | 246 |
| 10 | Black heterosexual women | 14374 | 14353 |
| 11 | Black gay/lesbian women | 438 | 438 |
| 12 | Black bisexual women | 1203 | 1193 |
| 13 | Hispanic/Latine heterosexual men | 16976 | 16954 |
| 14 | Hispanic/Latine gay men | 528 | 528 |
| 15 | Hispanic/Latine bisexual men | 466 | 465 |
| 16 | Hispanic/Latine heterosexual women | 19004 | 18982 |
| 17 | Hispanic/Latine gay/lesbian women | 436 | 434 |
| 18 | Hispanic/Latine bisexual women | 1523 | 1515 |
| 19 | Asian heterosexual men | 5113 | 5109 |
| 20 | Asian gay men | 124 | 124 |
| 21 | Asian bisexual men | 130 | 130 |
| 22 | Asian heterosexual women | 5300 | 5299 |
| 23 | Asian gay/lesbian women | 67 | 67 |
| 24 | Asian bisexual women | 280 | 279 |
| 25 | NAAN heterosexual men | 1401 | 1401 |
| 26 | NAAN gay men | 37 | 37 |
| 27 | NAAN bisexual men | 46 | 46 |
| 28 | NAAN heterosexual women | 1498 | 1496 |
| 29 | NAAN gay/lesbian women | 34 | 34 |
| 30 | NAAN bisexual women | 146 | 145 |
| 31 | NHPI heterosexual men | 524 | 524 |
| 32 | NHPI gay men | 18 | 18 |
| 33 | NHPI bisexual men | 20 | 20 |
| 34 | NHPI heterosexual women | 509 | 508 |
| 35 | NHPI gay/lesbian women | 16 | 16 |
| 36 | NHPI bisexual women | 32 | 32 |
| 37 | Multiracial heterosexual men | 3359 | 3350 |
| 38 | Multiracial gay men | 105 | 105 |
| 39 | Multiracial bisexual men | 144 | 143 |
| 40 | Multiracial heterosexual women | 3393 | 3379 |
| 41 | Multiracial gay/lesbian women | 145 | 145 |
| 42 | Multiracial bisexual women | 628 | 627 |

*Notes:* MDE = Major depressive episode. NAAN = Native American or Alaska Native. NHPI = Native Hawaiian or Pacific Islander.
